# Bailout cardiac surgery in patients undergoing transcatheter aortic valve replacement: a comprehensive analysis of post-marketing safety reports

**DOI:** 10.64898/2026.08.25.26361376

**Authors:** Salvatore Giordano, Nicola Corcione, Alberto Morello, Michele Cimmino, Michele Albanese, Paolo Ferraro, Giovanni Vecchione, Ignacio Amat-Santos, Arturo Giordano, Giuseppe Biondi-Zoccai

**Affiliations:** Cardiology Department, University Clinical Hospital Valladolid Valladolid, Spain; Cardiovascular Interventional Unit, Pineta Grande Hospital, Castel Volturno, Caserta, Italy; Cardiac Surgery Unit, Pineta Grande Hospital, Castel Volturno, Caserta, Italy; Centro de Investigación Biomédica en Red - Enfermedades Cardiovasculares (CIBERCV), Instituto de Salud Carlos III Madrid, Spain; Department of Medical-Surgical Sciences and Biotechnologies, Sapienza University of Rome, Latina, Italy; IRCCS Maria Cecilia Hospital – GVM Care & Research, Cotignola, Italy

**Keywords:** Aortic stenosis, bailout surgery, cardiac surgery, MAUDE, safety, transcatheter aortic valve implantation, transcatheter aortic valve replacement

## Abstract

**Background:** Bailout cardiac surgery during transcatheter aortic valve replacement (TAVR) is uncommon but remains associated with substantial morbidity and mortality. Although registries have described its incidence and major causes, they often provide limited detail regarding device-related failure mechanisms, attempted transcatheter rescue, and the clinical pathway leading to surgical conversion. We aimed at analyzing post-marketing safety reports from the U.S. Food and Drug Administration (FDA) Manufacturer and User Facility Device Experience (MAUDE) database to characterize the mechanisms, management strategies, and reported outcomes of bailout surgery during or shortly after TAVR.

**Methods:** We retrospectively analyzed FDA MAUDE reports received from July 1, 2016, through June 30, 2026. Eligible reports described unplanned urgent or emergent open cardiac surgery during or immediately after TAVR. Candidate reports were screened, adjudicated, and deduplicated at the clinical-event level. Events were classified by precipitating complication, transcatheter rescue, operative pathway, and reported outcome. Associations were evaluated using permutation tests, Fisher exact tests with Benjamini–Hochberg correction, adjusted regression models, and sensitivity analyses.

**Results:** After screening 43,239 initial reports, we identified 376 bailout-surgery events, with survival status was documented in 254, including 104 deaths and 150 survivors, corresponding to 40.9% reported mortality. Valve embolization, migration, or malposition was the most frequent complication phenotype (32.4%), whereas ventricular perforation or laceration was associated with the highest mortality (74.1%; OR, 4.86; 95% CI, 1.97–11.99). Mortality differed across complication phenotypes (*p*<0.001) and operative pathways (*p*<0.001), but not across transcatheter rescue pathways (*p*=0.355). Valve explantation with SAVR was associated with lower reported mortality (18.9%; OR, 0.29; 95% CI, 0.12–0.69), whereas unspecified surgery or access/support alone was associated with higher mortality (56.9%; OR, 3.04; 95% CI, 1.80–5.12). Ancillary analyses identified potential platform-specific differences in complication and management patterns, while bailout timing was not independently associated with mortality after adjustment.

**Conclusions:** In this MAUDE analysis, bailout cardiac surgery after TAVR was most commonly precipitated by valve embolization, migration, or malposition, whereas ventricular perforation or laceration was associated with the highest reported mortality. Outcomes differed across complication and operative pathways but not across transcatheter rescue strategies or bailout timing after adjustment. These findings identify clinically relevant post-marketing safety signals but should not be interpreted as incidence estimates, comparative device risks, or causal treatment effects.

## Introduction

Severe aortic stenosis is a progressive and life-threatening valvular disorder for which aortic valve replacement remains the definitive treatment.(1) Transcatheter aortic valve replacement (TAVR), initially developed for patients considered inoperable or at high surgical risk, has progressively expanded to intermediate- and low-risk populations.(2) Contemporary practice now includes younger patients and increasingly complex scenarios, such as bicuspid valves, heavily calcified anatomy, valve-in-valve procedures, and alternative-access interventions.(3) In parallel, advances in transcatheter heart-valve design, delivery-system profiles, multidetector computed-tomography planning, implantation techniques, and multidisciplinary Heart Team pathways have substantially improved procedural safety.(4) Nevertheless, the broader application of TAVR across heterogeneous anatomical and clinical settings continues to expose patients to rare but potentially catastrophic complications requiring immediate rescue.(5)

Bailout cardiac surgery refers to unplanned emergency conversion to open cardiac surgery during or shortly after transcatheter aortic valve replacement, typically in response to hemodynamic collapse or a life-threatening complication not amenable to percutaneous management.(6) While quite rare, it carries substantial perioperative morbidity and mortality and may require sternotomy or thoracotomy, with or without cardiopulmonary bypass.(7) The principal triggers include annular or left ventricular outflow tract rupture, ventricular perforation, aortic dissection or rupture, valve embolization or malposition, acute prosthetic dysfunction, coronary obstruction, mitral apparatus injury, and unretrievable device components.(8-9) These events often result from a complex interaction among adverse anatomy, heavy calcification, prosthesis sizing or positioning, guidewire or balloon-related trauma, delivery-system issues, and procedural technique.(10-12)

Although transcatheter rescue strategies such as pericardiocentesis, valve-in-valve implantation, snaring, coronary or covered stenting, and mechanical circulatory support may avert surgery in selected cases, bailout cardiac surgery remains necessary when catheter-based rescue is infeasible, unsuccessful, or delayed.(11-13) Outcomes in these catastrophic scenarios may depend on rapid recognition, immediate multidisciplinary coordination, and timely access to cardiac surgery, perfusion support, and cardiopulmonary bypass, thereby sustaining the debate over the safety of TAVR at centers without on-site surgery.(7,14) Existing registries and institutional series have defined the incidence, major causes, and outcomes of surgical conversion, but they often provide limited detail regarding device behavior, the sequence of rescue attempts, and the reasons for failure-to-rescue.(7,15-17)

The U.S. Food and Drug Administration (FDA) Manufacturer and User Facility Device Experience (MAUDE) database offers a complementary narrative-rich source for characterizing these pathways, although its inherent limitations require that findings be interpreted as descriptive post-marketing safety signals rather than estimates of incidence or comparative device risk.(18-21) We thus aimed at leveraging the MAUDE dataset to characterize from the unique perspective of safety reporting bailout cardiac surgery for TAVR.

## Methods

We conducted a retrospective, descriptive post-marketing surveillance study of reports in the MAUDE database from July 1, 2016, through June 30, 2026.

TAVR systems were identified using a prespecified combination of manufacturers and device names (eg Abbott and Navitor). Narratives were searched for terms indicating emergency conversion, including sternotomy, cardiopulmonary bypass, open-heart surgery, explantation, and surgical bailout. The primary cohort comprised unplanned urgent or emergent open cardiac surgery during TAVR or immediately postprocedurally. A broader sensitivity cohort included early-delayed surgery during the index hospitalization. Planned operations, isolated peripheral vascular repairs, late endocarditis or structural deterioration, aggregate or literature reports, and events lacking adequate evidence of qualifying surgery or timing were excluded.

Two trained reviewers adjudicated candidate reports and resolved disagreements by consensus. Duplicate, follow-up, amended, or companion submissions were consolidated using report identifiers, event dates, device characteristics, and narrative concordance. Extracted variables included valve platform, timing and procedural stage, precipitating complication, attempted transcatheter rescue, operative intervention, perioperative support, and reported outcome. Prespecified taxonomies classified complications and reduced management sequences to mutually exclusive dominant rescue and operative pathways.

Categorical variables were summarized as counts and percentages, and mortality analyses were restricted to events with documented survival status.(22) Associations between complication, rescue, or operative categories and mortality were assessed using 20,000-permutation global tests with Cramér’s V and category-versus-all-other Fisher exact tests, with odds ratios and 95% confidence intervals.(23) Sparse categories were collapsed only for global testing, and *p* values were adjusted using the Benjamini–Hochberg false-discovery-rate procedure.(24) Ancillary analyses compared self-expanding and balloon-expandable platforms using 100,000-permutation fixed-margin tests; mechanically expanded/repositionable valves were retained descriptively but excluded from formal comparisons. Statistical significance was set at the 2-tailed 0.05 level. Computations were performed with Python 3.14.6 (Python Software Foundation, Beaverton, OR, USA)

## Results

From a total of 43,239 initial reports, screening yielded a dataset of 624 candidate reports, all of which underwent report-level screening and clinical adjudication. After application of the prespecified eligibility and timing criteria, 248 reports (39.7%) were excluded from the primary cohort, leaving 376 qualifying events (60.3%); 14 additional events were retained only in the broad cohort, yielding 390 events for sensitivity analyses. The most frequent reasons for non-inclusion were late endocarditis or structural deterioration outside the bailout window (58 reports), insufficient evidence of qualifying bailout surgery or timing (58), and aggregate or literature-based reporting rather than a unique patient event (38). A complete clinical sequence encompassing the precipitating mechanism, rescue attempt, operative intervention, and outcome was available for 128 primary-cohort events (34.0%), whereas 248 (66.0%) lacked at least one of these elements. Outcome status was reported for 254 events (67.6%) and was unavailable for 122 (32.4%); mortality analyses were therefore restricted to events with known outcomes.

The primary cohort comprised 376 unique clinical events involving bailout cardiac surgery during or shortly after TAVR, reconstructed according to the sequence of precipitating complication, attempted transcatheter rescue, operative pathway, and reported outcome. Clinical outcome was documented in 254 events (67.6%), including 104 deaths and 150 survivors, corresponding to a reported mortality of 40.9% among events with known outcomes; outcome information was unavailable for the remaining 122 events (32.4%) (Figure 1). Valve embolization, migration, or malposition was the most frequent complication phenotype (122 events, 32.4%; Table 1). A valve-directed transcatheter rescue strategy was documented in 239 events (63.6%), including balloon intervention in 84 (22.3%), second-valve or valve-in-valve implantation in 80 (21.3%), and snare-based retrieval or repositioning in 75 (19.9%). The most frequent operative categories were an unspecified operation, surgical access, or support without a clearly documented definitive repair (155 events, 41.2%); valve explantation without documented surgical aortic valve replacement (SAVR; 109, 29.0%); valve explantation with SAVR (54, 14.4%); and SAVR without documented explantation (39, 10.4%; Table 2). Associations with reported mortality were evaluated among events with known outcomes using permutation-based global tests and category-versus-all-other Fisher exact tests, with Benjamini–Hochberg correction for multiple comparisons.

**Table 1.** Precipitating complication phenotypes and reported outcomes.

| Complication phenotype | All reports, | Known outcomes, | Deaths, | Survivors, | Mortality, | OR for death | Fisher | BH-FDR |
| --- | --- | --- | --- | --- | --- | --- | --- | --- |
|  | n (%) | n (%) | n | n | % | (95% CI) | p | q |
| Valve embolization/migration/malposition | 122 (32.4) | 79 (64.8) | 20 | 59 | 25.3 | 0.37 (0.20–0.66) | <0.001 | 0.004 |
| Other/indeterminate | 74 (19.7) | 48 (64.9) | 18 | 30 | 37.5 | 0.84 (0.44–1.60) | 0.628 | 0.921 |
| Aortic dissection/rupture | 55 (14.6) | 44 (80.0) | 19 | 25 | 43.2 | 1.12 (0.58–2.16) | 0.739 | 0.921 |
| Annular/LVOT/aortic-root rupture | 37 (9.8) | 28 (75.7) | 14 | 14 | 50.0 | 1.51 (0.69–3.32) | 0.315 | 0.841 |
| Ventricular perforation/laceration | 36 (9.6) | 27 (75.0) | 20 | 7 | 74.1 | 4.86 (1.97–11.99) | <0.001 | 0.002 |
| Tamponade/effusion—source unclear | 33 (8.8) | 18 (54.5) | 8 | 10 | 44.4 | 1.17 (0.44–3.06) | 0.806 | 0.921 |
| Coronary obstruction/occlusion | 10 (2.7) | 7 (70.0) | 3 | 4 | 42.9 | 1.08 (0.24–4.95) | NE | NE |
| Acute prosthetic dysfunction/regurgitation/PVL | 8 (2.1) | 3 (37.5) | 2 | 1 | 66.7 | 2.92 (0.26–32.65) | 0.569 | 0.921 |
| Device/component entrapment | 1 (0.3) | 0 (0.0) | — | — | — | — | — | — |

Data are n (%) unless otherwise indicated. Percentages in “All reports” use the full cohort (N=376); percentages in “Known outcomes” are category-specific outcome-reporting rates. Mortality analyses included only reports with a documented outcome (104 deaths and 150 survivors; n=254); 122 reports had no documented outcome. Odds ratios compare each category with all other complication categories. Fisher exact p values were adjusted using the Benjamini-Hochberg false-discovery-rate procedure. The global association between collapsed complication phenotype and reported mortality was significant (20,000-permutation p=0.0009; Cramér’s V=0.294). Sparse categories were collapsed only for the global test. Bold q values indicate q<0.05. Abbreviations: BH-FDR, Benjamini-Hochberg false-discovery rate; CI, confidence interval; LVOT, left ventricular outflow tract; NE, not estimable; OR, odds ratio; PVL, paravalvular leak.

**Table 2.**
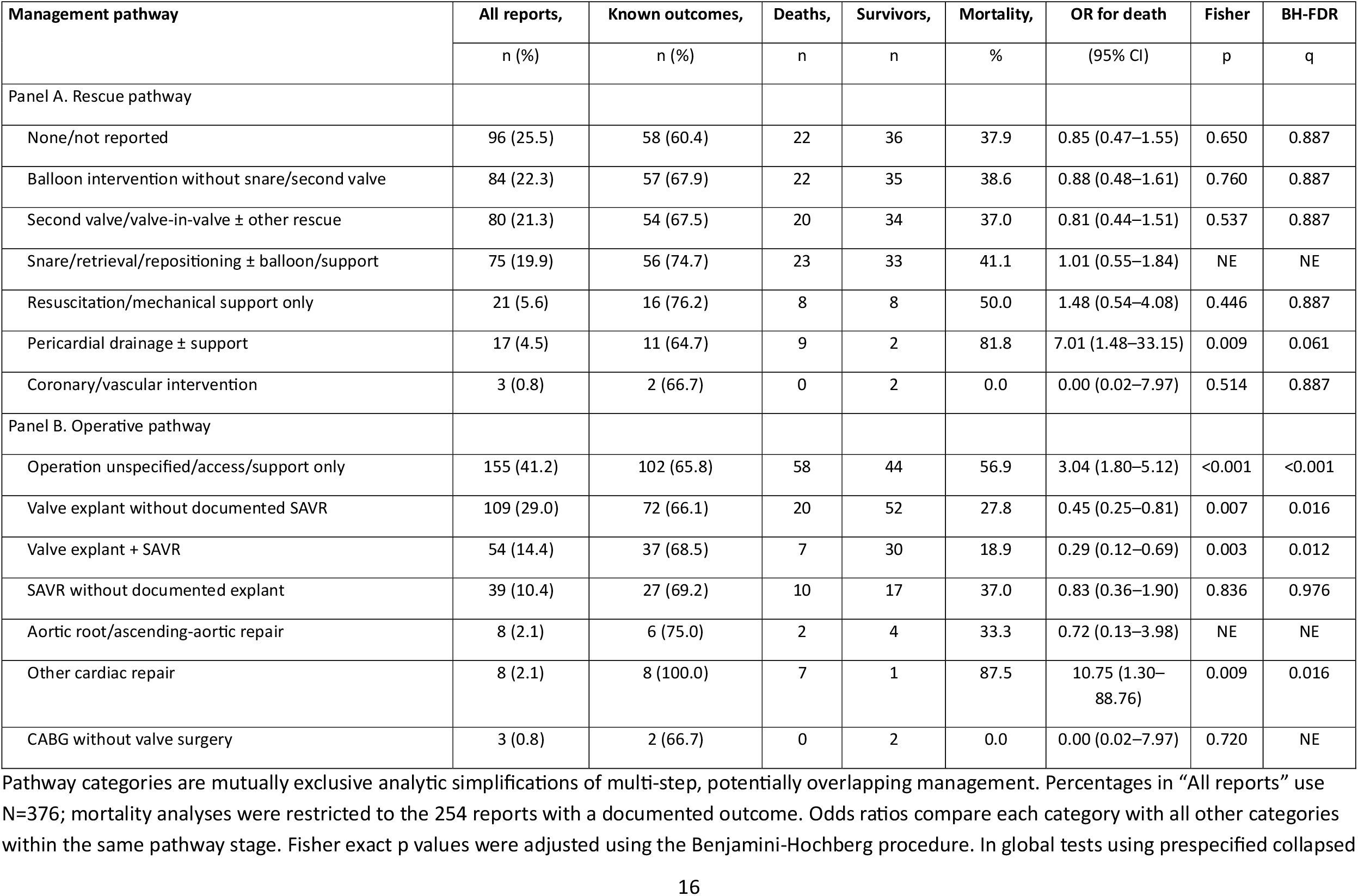
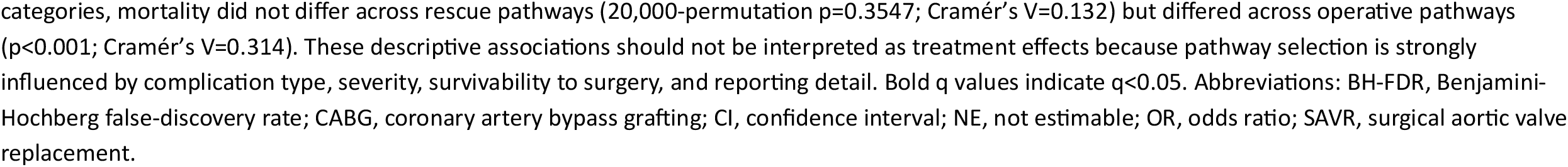
Transcatheter rescue and operative pathways with reported outcomes.

| Management pathway | All reports, | Known outcomes, | Deaths, | Survivors, | Mortality, | OR for death | Fisher | BH-FDR |
| --- | --- | --- | --- | --- | --- | --- | --- | --- |
|  | n (%) | n (%) | n | n | % | (95% CI) | p | q |
| Panel A. Rescue pathway |  |  |  |  |  |  |  |  |
| None/not reported | 96 (25.5) | 58 (60.4) | 22 | 36 | 37.9 | 0.85 (0.47–1.55) | 0.650 | 0.887 |
| Balloon intervention without snare/second valve | 84 (22.3) | 57 (67.9) | 22 | 35 | 38.6 | 0.88 (0.48–1.61) | 0.760 | 0.887 |
| Second valve/valve-in-valve ± other rescue | 80 (21.3) | 54 (67.5) | 20 | 34 | 37.0 | 0.81 (0.44–1.51) | 0.537 | 0.887 |
| Snare/retrieval/repositioning ± balloon/support | 75 (19.9) | 56 (74.7) | 23 | 33 | 41.1 | 1.01 (0.55–1.84) | NE | NE |
| Resuscitation/mechanical support only | 21 (5.6) | 16 (76.2) | 8 | 8 | 50.0 | 1.48 (0.54–4.08) | 0.446 | 0.887 |
| Pericardial drainage ± support | 17 (4.5) | 11 (64.7) | 9 | 2 | 81.8 | 7.01 (1.48–33.15) | 0.009 | 0.061 |
| Coronary/vascular intervention | 3 (0.8) | 2 (66.7) | 0 | 2 | 0.0 | 0.00 (0.02–7.97) | 0.514 | 0.887 |
| Panel B. Operative pathway |  |  |  |  |  |  |  |  |
| Operation unspecified/access/support only | 155 (41.2) | 102 (65.8) | 58 | 44 | 56.9 | 3.04 (1.80–5.12) | <0.001 | <0.001 |
| Valve explant without documented SAVR | 109 (29.0) | 72 (66.1) | 20 | 52 | 27.8 | 0.45 (0.25–0.81) | 0.007 | 0.016 |
| Valve explant + SAVR | 54 (14.4) | 37 (68.5) | 7 | 30 | 18.9 | 0.29 (0.12–0.69) | 0.003 | 0.012 |
| SAVR without documented explant | 39 (10.4) | 27 (69.2) | 10 | 17 | 37.0 | 0.83 (0.36–1.90) | 0.836 | 0.976 |
| Aortic root/ascending-aortic repair | 8 (2.1) | 6 (75.0) | 2 | 4 | 33.3 | 0.72 (0.13–3.98) | NE | NE |
| Other cardiac repair | 8 (2.1) | 8 (100.0) | 7 | 1 | 87.5 | 10.75 (1.30–88.76) | 0.009 | 0.016 |
| CABG without valve surgery | 3 (0.8) | 2 (66.7) | 0 | 2 | 0.0 | 0.00 (0.02–7.97) | 0.720 | NE |

Pathway categories are mutually exclusive analytic simplifications of multi-step, potentially overlapping management. Percentages in “All reports” use N=376; mortality analyses were restricted to the 254 reports with a documented outcome. Odds ratios compare each category with all other categories within the same pathway stage. Fisher exact p values were adjusted using the Benjamini-Hochberg procedure. In global tests using prespecified collapsed
categories, mortality did not differ across rescue pathways (20,000-permutation $p=0.3547$ ; Cramér's $V=0.132$ ) but differed across operative pathways ( $p<0.001$ ; Cramér's $V=0.314$ ). These descriptive associations should not be interpreted as treatment effects because pathway selection is strongly influenced by complication type, severity, survivability to surgery, and reporting detail. Bold q values indicate $q<0.05$ . Abbreviations: BH-FDR, Benjamini-Hochberg false-discovery rate; CABG, coronary artery bypass grafting; CI, confidence interval; NE, not estimable; OR, odds ratio; SAVR, surgical aortic valve replacement.

**Figure 1.**
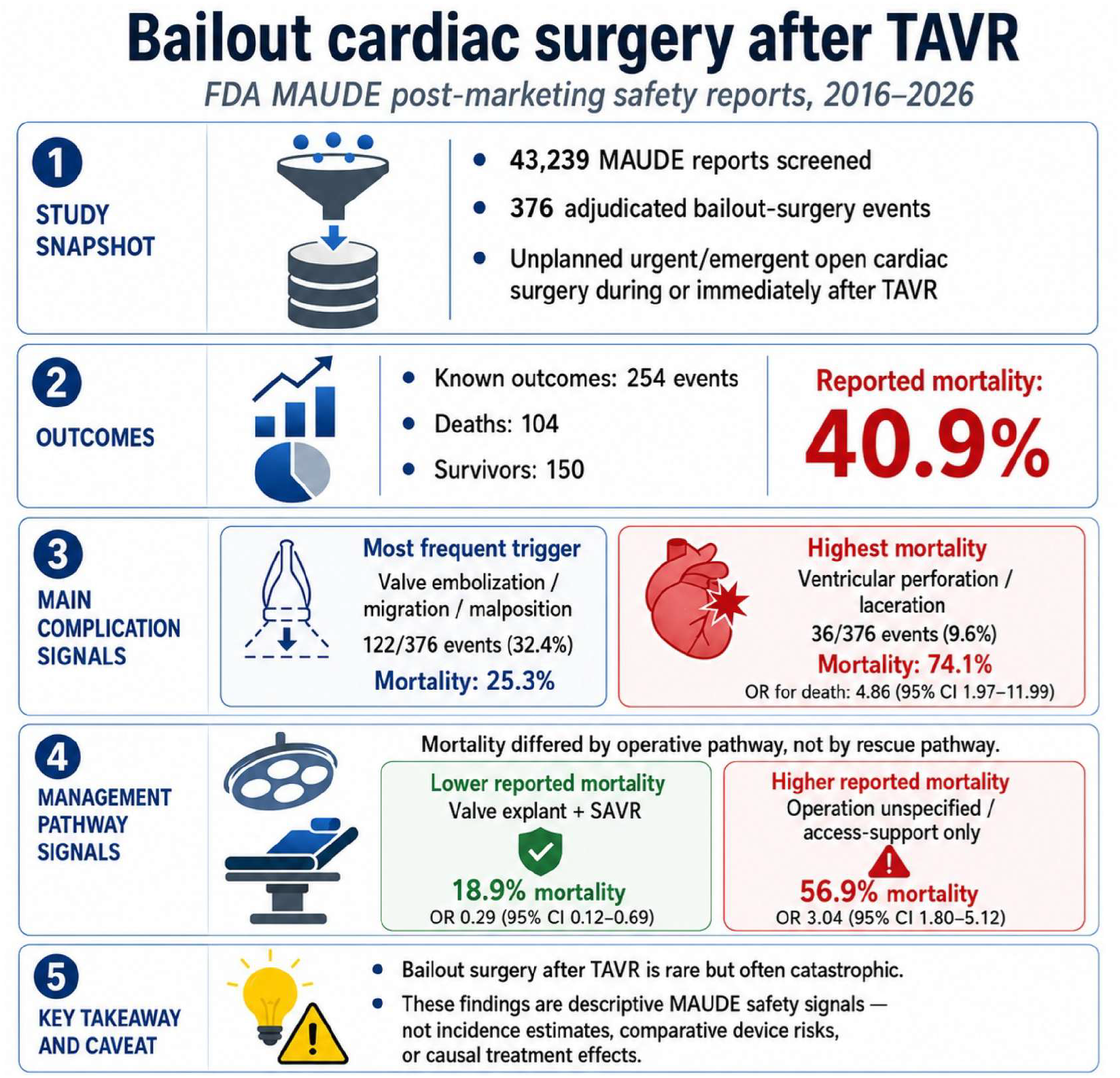
Mechanisms, management, and reported outcomes of bailout cardiac surgery in patients undergoing transcatheter aortic valve replacement (TAVR) in the FDA MAUDE database. CI, confidence interval; FDA, U.S. Food and Drug Administration; MAUDE, Manufacturer and User Facility Device Experience; OR, odds ratio.

As shown in Table 1, mortality distributions differed significantly across complication phenotypes (20,000-permutation *p*<0.001; Cramér’s V=0.294). Ventricular perforation or laceration was associated with the highest mortality among the principal complication categories, with 20 deaths among 27 events with known outcomes (74.1%; odds ratio [OR], 4.86; 95% confidence interval [CI], 1.97–11.99; false-discovery-rate-adjusted *q*<0.001). Conversely, valve embolization, migration, or malposition was associated with lower reported mortality, with 20 deaths among 79 events with known outcomes (25.3%; OR, 0.37; 95% CI, 0.20–0.66; *q*=0.004). In Table 2, mortality did not differ significantly across rescue pathways in the global analysis (*p*=0.355; Cramér’s V=0.132); although pericardial drainage with or without supportive measures was associated with 9 deaths among 11 known outcomes (81.8%; OR, 7.01; 95% CI, 1.48–33.15), this association did not remain significant after multiple-testing correction (*q*=0.061). In contrast, mortality differed across operative pathways (*p*<0.001; Cramér’s V=0.314). An unspecified operation or surgical access/support alone was associated with higher reported mortality (58/102, 56.9%; OR, 3.04; 95% CI, 1.80–5.12; *q*<0.001), as was other cardiac repair, although the latter estimate was based on only eight events (7/8 deaths, 87.5%; OR, 10.75; 95% CI, 1.30–88.76; *q*=0.016). Lower mortality was observed for valve explantation with SAVR (7/37, 18.9%; OR, 0.29; 95% CI, 0.12–0.69; *q*=0.012) and valve explantation without documented SAVR (20/72, 27.8%; OR, 0.45; 95% CI, 0.25–0.81; *q*=0.016). These associations are descriptive and should not be interpreted as treatment effects because management selection was influenced by complication type, clinical severity, survivability to surgery, and the completeness of the submitted narratives.

In ancillary analyses, complication profiles differed between self-expanding and balloon-expandable valve reports (permutation χ^2^=31.46, *p*<0.001; Cramér’s V=0.292; Table 1S), predominantly because annular, left ventricular outflow tract, or aortic-root rupture was more frequently represented among balloon-expandable than self-expanding cases (13/37 [35.1%] vs 24/333 [7.2%]; OR, 6.97; 95% CI, 3.16–15.40; *q*<0.001). After adjustment for complication phenotype and reporting era, a transcatheter rescue attempt was less frequently documented in balloon-expandable cases (29.7% vs 79.0%; adjusted OR, 0.09; 95% CI, 0.04–0.21; *p*<0.001), whereas SAVR was more frequently reported (43.2% vs 22.5%; adjusted OR, 2.60; 95% CI, 1.21–5.57; *p*=0.014); valve explantation and cardiopulmonary bypass did not differ significantly after adjustment (Table 2S). Complication distributions were similar across the complete 2016–2019, 2020–2022, and 2023–2026 eras (permutation *p*=0.813), although the relative representation of balloon-expandable reports increased with event year (OR per year, 1.21; 95% CI, 1.00–1.46; *p*=0.045), and median event-to-FDA-receipt delay decreased from 22.5 days in 2016–2019 to 17.0 days in 2020–2022 and 13.5 days in 2023–2026 (Kruskal–Wallis H=30.37; *p*<0.001). Bailout occurred intraprocedurally in 218 events, immediately postprocedurally with unquantified timing in 109, and within 24 hours in 49; corresponding mortality among events with documented outcomes was 35.5%, 50.0%, and 47.2%, respectively. However, neither the unadjusted global comparison (*p*=0.099) nor the analysis adjusted for complication phenotype and valve design supported an independent association between timing and mortality (global *p*=0.175); compared with intraprocedural bailout, adjusted risk ratios were 1.31 (95% CI, 0.96–1.80) for immediate events with unquantified timing and 1.33 (95% CI, 0.88–2.02) for events within 24 hours. Timing was also not associated with the principal operative procedures, and nominal differences in rescue strategies did not remain significant after false-discovery-rate correction. Restriction to 352 independently classified events yielded materially unchanged findings (Table 3S).

## Discussion

This post-marketing surveillance analysis confirmed bailout surgery for TAVR as a rare complication: assuming a total TAVR volume between 2016 and 2026 of 900,000 cases in the U.S. and systematic reporting in MAUDE, an informed estimate of incidence of bailout surgery for TAVR is 0.4% during the last 10 years.(25) Among reported cases, valve embolization, migration, or malposition were the most frequently reported complications precipitating bailout surgery, whereas ventricular perforation or laceration was associated with the highest reported mortality. By reconstructing the sequence from precipitating complication to transcatheter rescue, operative intervention, and outcome, the study provides a detailed perspective of bailout pathways that is not typically available in structured registries. These narrative-rich MAUDE reports therefore complement registry and administrative data by clarifying device-related mechanisms, rescue attempts, and the surgical procedures ultimately required.(7,14) Among events with documented survival status, mortality was 40.9%, underscoring the substantial clinical severity of surgical bailout while remaining interpretable only as a report-level post-marketing safety signal.

The precipitating complications leading to bailout cardiac surgery encompassed a broad spectrum of catastrophic structural and device-related events, underscoring the heterogeneous mechanisms through which TAVR may require emergent conversion.(15) Valve embolization, migration, or malposition was the most frequently represented phenotype, likely reflecting both its procedural visibility and the detailed reporting of device-related events in MAUDE.(26) Despite its quite low frequency, this category was associated with comparatively lower reported mortality, possibly because early recognition and transcatheter stabilization may permit a more controlled transition to definitive surgery. In contrast, ventricular perforation or laceration carried the highest mortality, consistent with the rapid hemodynamic deterioration, tamponade, and limited rescue interval characteristic of these injuries.(28) Annular, left ventricular outflow tract, aortic-root, aortic-wall, coronary, and acute prosthetic complications were less frequently represented but remained clinically important because they often required complex repair beyond isolated valve replacement.(29)

Transcatheter rescue was frequently attempted before surgical conversion, most commonly through balloon intervention, second-valve implantation, or snare-based retrieval and repositioning. However, reported mortality did not differ significantly across rescue pathways, suggesting that the selected strategy largely reflected the underlying complication, anatomy, and hemodynamic condition rather than an independently modifiable determinant of outcome.(7) The high mortality observed after pericardial drainage or mechanical support should therefore be interpreted primarily as a marker of severe structural injury and circulatory collapse.(12) Operative management was similarly heterogeneous, ranging from surgical access or support alone to valve explantation, SAVR, coronary bypass, ventricular repair, and complex aortic reconstruction. Higher mortality after an unspecified operation or access/support alone may indicate extreme instability, incomplete procedural documentation, or death before definitive repair, whereas the lower mortality observed after valve explantation with or without SAVR may partly reflect survivorship and treatment-selection bias.(15) Accordingly, the observed differences across operative pathways should be regarded as descriptive associations rather than evidence of the comparative effectiveness of specific rescue or surgical strategies.

Ancillary analyses showed that complication profiles differed by valve platform, driven primarily by the greater representation of annular, LVOT, or aortic-root rupture among balloon-expandable than self-expanding reports. After adjustment for complication phenotype and reporting era, transcatheter rescue was less frequently documented with balloon-expandable valves, whereas SAVR was more frequently reported. Event-to-FDA-receipt delays shortened significantly across over the years, suggesting progressively faster post-marketing reporting. Although mortality was numerically higher when bailout occurred after the procedure, timing was not independently associated with death, reinforcing the importance of continuous postprocedural surveillance and immediate access to coordinated interventional and surgical rescue.(30)

These findings support systematic preprocedural bailout planning, multidisciplinary Heart Team assessment, and immediate access to cardiac surgical, perfusion, and mechanical-support expertise during TAVR.(31) However, the MAUDE database is subject to selective and incomplete reporting, missing outcomes, potential residual duplicate events, and the absence of procedural denominators.(32-33) Accordingly, the present results should not be interpreted as incidence estimates, comparative device risks, temporal safety trends, or causal treatment effects.(34) Future studies should integrate narrative post-marketing surveillance with linked registries, standardized event adjudication, time-to-intervention metrics, and longitudinal outcome assessment.

In conclusion, valve embolization, migration, or malposition was the most frequently reported trigger for bailout surgery after TAVR, whereas ventricular perforation or laceration carried the highest mortality. Event-pathway reconstruction highlighted the heterogeneity and severity of these emergencies and the need for immediate multidisciplinary surgical support. Given MAUDE’s absent denominators, incomplete reporting, and potential duplication, these findings should be considered descriptive safety signals requiring registry-based confirmation.

## Data Availability

The data used for this study can be requested from the corresponding author for research purposes.

## Acknowledgements

This manuscript was drafted with the assistance of artificial intelligence tools, such as ChatGPT (OpenAI, San Francisco, CA, USA), in keeping with established best practices (Biondi-Zoccai G, editor. ChatGPT for Medical Research. Torino: Edizioni Minerva Medica; 2024). The final content, including all conclusions and opinions, has been thoroughly revised, edited, and approved by the authors. The authors take full responsibility for the integrity and accuracy of the work and retain full credit for all intellectual contributions. Compliance with ethical standards and guidelines for the use of artificial intelligence in research has been ensured.

